# Evaluating self-triage accuracy of laypeople, symptom-assessment apps, and large language models: A framework for case vignette development using a representative design approach (RepVig)

**DOI:** 10.1101/2024.04.02.24305193

**Authors:** Marvin Kopka, Hendrik Napierala, Martin Privoznik, Desislava Sapunova, Sizhuo Zhang, Markus A. Feufel

## Abstract

Most studies evaluating symptom-assessment applications (SAAs) rely on a common set of case vignettes that are authored by clinicians and devoid of context, which may be representative of clinical settings but not of situations where patients use SAAs. Assuming the use case of self-triage, we used representative design principles to sample case vignettes from online platforms where patients describe their symptoms to obtain professional advice and compared triage performance of laypeople, SAAs, and Large Language Models (LLMs) on representative versus standard vignettes. We found performance differences in all three groups depending on vignette type (OR = 1.27 to 3.41, p < .001 to .035) and changed rankings of best-performing SAAs and LLMs. Based on these results, we argue that our representative vignette sampling approach (that we call the RepVig Framework) should replace the practice of using a fixed vignette set as standard for SAA evaluation studies.

## Introduction

Symptom-assessment applications (SAAs) are digital health tools that assist medical laypeople in self-diagnosing and determining whether and where to seek health care (self-triage)^1,2^. Whereas some research in this domain focuses on how SAAs impact health systems or on how to improve their usability and user experience^2–7^, a significant portion of studies investigates the accuracy of SAAs^8,9^. This line of research started with Semigran et al. in 2015,^10^ who developed 45 case vignettes to systematically test and compare the accuracy of SAAs. These vignettes were developed by clinicians, drawing from a variety of medical resources including textbooks for medical education. Subsequently, numerous studies have adopted their methodology and/or vignettes. Some authors have used the same set of vignettes,^11–13^ whereas others have developed their own set, either building on the original vignettes or employing a similar approach to create them.^14–16^ Over time, these case vignettes have faced various criticisms: in addition to procedural criticism (e.g., that it matters who inputs vignettes into SAAs and that interrater reliabilities should be calculated^17^), most criticism refers to the content of the case vignettes (e.g., the symptoms and symptom clusters covered in the cases)^8^ or to how they were created (e.g., the vignettes are usually created by clinicians, who may describe symptoms differently than patients, and are often fictitious rather than based on real cases)^18^.

To mitigate these issues, SAAs were tested with cases that more closely resemble real-world situations and actual patients. For instance, Yu et al.^19^ and Berry et al.^20^ utilized patient data from Emergency Department (ED) presentations for their vignettes. This method enhances external validity – understood as the applicability of findings to real-world scenarios^21^ – yet its representativeness is still limited with respect to the primary purpose of SAAs: aiding medical laypeople experiencing acute symptoms deciding if or where to seek care^22^. That is, these vignettes include problems of people who chose to visit an ED, a group that may not fully represent the wider array of SAA users who (rightly or wrongly) decided against going to the ED in the first place. Additionally, the case development was biased, as clinicians re-created these vignettes based on their documentation, who may have inadvertently filtered information or phrased the vignettes differently than laypeople using an SAA would have done^18^. Assuming the use case of self-triage, previous studies thus did not yet generate a representative set of vignettes. Results of SAA audit studies that are based on such stimuli might not be generalizable to laypeople’s self-triage with SAA in the real world.

The conceptual challenge of generating a representative set of stimuli was recognized in decision-making research decades ago, particularly in the field of ecological psychology rooted in Egon Brunswik’s probabilistic functionalism^23^. In his framework, decision-makers infer unobservable conditions (“latent variables” or a triage level) from a set of observable information (“cues” or symptoms) based on characteristic correlations between certain cues and the latent variable (see Dhami et al.^21^ for an overview). Figure 1 implements these considerations for self-triage decisions.

**Figure 1.**
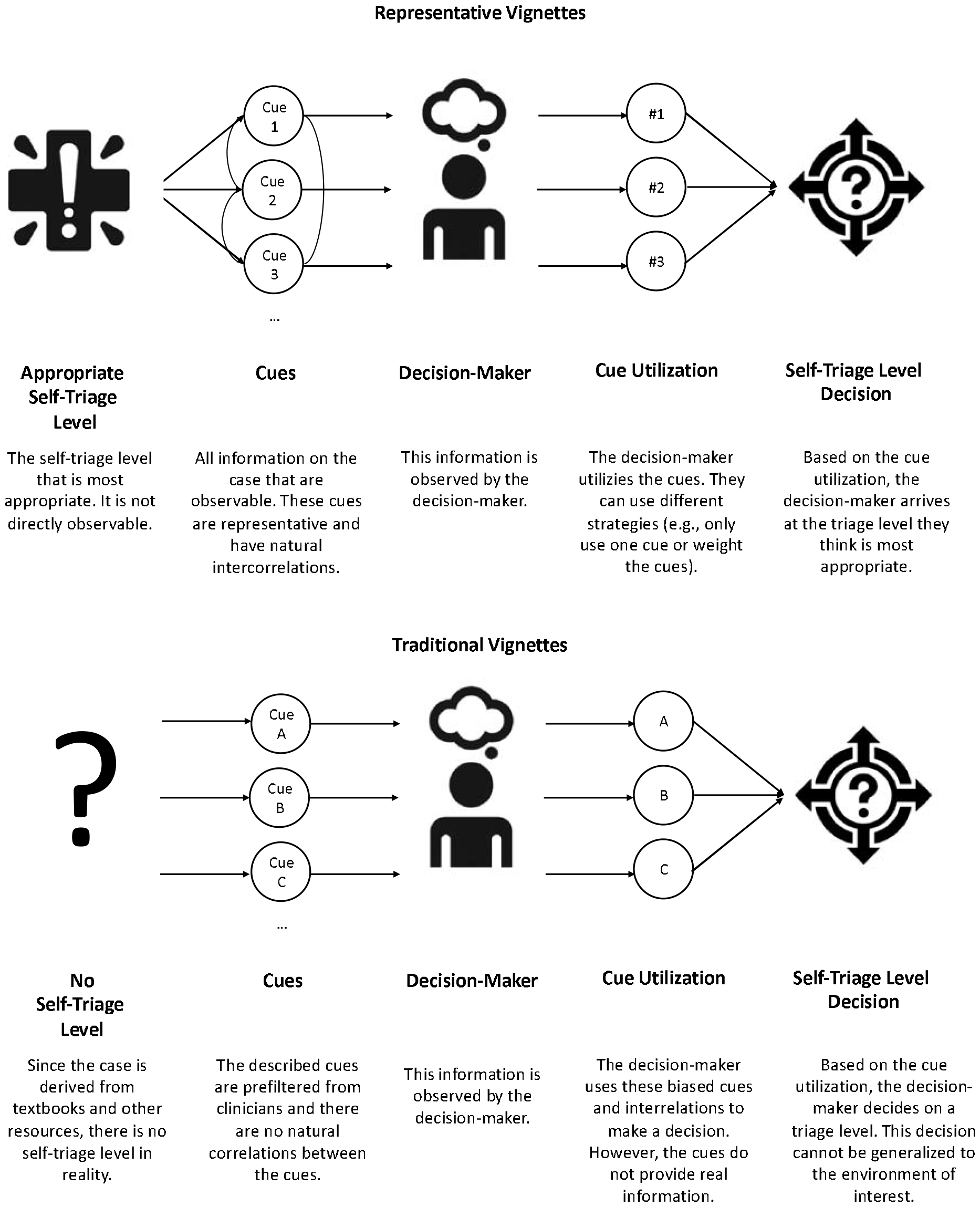
Self-triage lens model.

Based on these considerations, Brunswik coined the term *ecological validity* and proposed the concept of *representative design* as a method for sampling ecologically valid stimuli. Using representative design, stimuli (or vignettes) are sampled directly from situations to which the researcher would like to generalize, thereby trying to maintain the natural correlations between cues and the latent variable^23^. This approach contrasts starkly with the traditional *systematic design*, where parameters of the stimuli are systematically varied, which according to representative design may lead to biased cues and intercorrelations^21^. For example, a higher age might be associated with a higher risk for certain diseases, and systematically varying age tends to eliminate these associations. As a result, decision-makers might make different decisions when given systematically designed stimuli. Conversely, stimuli derived based on representative design help researchers induce and evaluate decision-making behavior that more readily generalizes to the environment of interest, in our case, from SAA audit studies to real-world self-triage performance of medical laypeople.

The power of representative design has been demonstrated in various fields such as pharmacy^24^, social psychology^25^, cognitive science^26^, human resources^27^, and public health^28^, showing that predictive power of results from experiments increases and these results better resemble real-world performance. Studies that compared representative stimuli with non-representative stimuli found significant differences between results and conclusions^25,27,29,30^. For example, ecologically valid stimuli tended to make phenomena identified with non-representative stimuli disappear or reverse the direction of effects^25^ and better predicted performance in field studies^27,29^.

To our knowledge, however, the representative design framework has thus far not been applied to study self-triage decisions and evaluate SAA performance. Our paper aims to bridge this research gap. Building on an application of the representative design approach to the context of self-triage decisions, we explore how a framework to generate representative vignettes (that we called the RepVig Framework) can be used to effectively generate new vignettes to assess self-triage capabilities. We compare results obtained from the vignettes we developed using the RepVig Framework with those developed with traditional approaches and test whether these results differ. We hypothesize that these two methods of vignette development will yield different results.

## Methods

### Ethical Considerations

This study was carried out in accordance with the declaration of Helsinki. Ethical approval was granted by the ethics committee of the Department of Psychology and Ergonomics (IPA) at Technische Universität Berlin (tracking number: AWB_KOP_2_230711). The study was preregistered in the WHO-accredited German Clinical Trials Register (ID: DRKS00032895). The methods and results are reported conforming to the STROBE reporting guideline^33^.

### Study Design

This study was designed as a prospective observational study examining the (self-)triage performance of medical laypeople, SAAs and Large Language Models (LLMs) based on two kinds of vignettes. We used a set of existing vignettes that have been used to evaluate triage performance of laypeople and SAAs in previous studies^34,35^. In addition, we aimed to develop a new vignette set following a representative design approach^23^ and the RepVig Framework, aiming for greater external validity in generalizing to self-triage decisions compared to prior studies. We investigated whether vignettes obtained using the RepVig framework yield different triage performance estimates compared to the traditional vignette sets. We gathered urgency assessments from medical laypeople and entered the vignettes in various SAAs and LLMs to determine standard metrics for triage-performance reporting^36,37^.

### Vignette Sets

Brunswik^23^ emphasized the importance of sampling stimuli (vignettes in this case) from a population of stimuli from the reference class. To gather such symptom descriptions from individuals who are (1) self-describing their symptoms and (2) in the process of deciding if and where to seek healthcare, we utilized the social media network Reddit, specifically the subreddit ‘r/AskDocs’. This forum allows people to post medical questions, which are then addressed by verified physicians on a voluntary basis. For our research, we used Reddit’s API to extract all new posts within a 14-day period, from June 16^th^ to June 29^th^ 2023.

Recognizing that the symptom distribution in these cases might differ from those typically entered into SAAs, we applied Brunswik’s second approach to achieving representative design, known as ‘canvassing’^21^. To this end, we established quotas based on symptom clusters commonly input into SAAs. We used data from Arellano Carmona et al.^38^ who examined symptom clusters based on the CDC’s National Ambulatory Medical Care Survey (and corresponding triage advice) that users entered into an SAA, and constructed these quotas to ensure that our vignette set is representative not only in terms of real-world symptom descriptions but also with respect to the symptom distributions that users tend to enter into SAAs. These quotas can be found in Table 1.

**Table 1.** Quotas used to sample vignettes that are representative of symptom clusters typically entered in SAA based on Arellano Carmona et al.^38^.

| Symptom Cluster | # of vignettes (%) |
| --- | --- |
| Musculoskeletal Pain | 5 (11%) |
| Joint Pain | 3 (7%) |
| Headache | 1 (2%) |
| Chest Pain | 1 (2%) |
| Other Pain | 5 (11%) |
| Gynecological | 5 (11%) |
| Tumors/lumps/masses | 4 (9%) |
| Edema | 2 (4%) |
| Skin issues | 2 (4%) |
| Gastrointestinal | 2 (4%) |
| Impaired sensations | 3 (7%) |
| Urinary Tract Problems | 3 (7%) |
| Upper Respiratory Symptoms | 1 (2%) |
| Other | 8 (18%) |
| Total Number | 45 (100%) |

To ensure that we include vignettes by people who are experiencing a situation in which they would consult an SAA (i.e., that they experienced new symptoms and sought advice on what to do), we applied the following exclusion criteria, omitting vignettes where authors:

‐ sought general information only,
‐ provided a picture to be understandable, as these are not typical SAA inputs,
‐ provided overly specific information (e.g., blood values that are typically not known to laypeople),
‐ discussed symptoms for which the individual had already consulted a medical professional,
‐ provided insufficient details (e.g., a single sentence stating foot pain) for self-triage,
‐ described symptoms experienced by someone else (with the exception of infants),
‐ did not describe acute symptoms (e.g., past symptoms that have already resolved)
‐ described symptoms already diagnosed by a medical professional
‐ exceeded 1,000 characters (which may overwhelm participants).

During the 14-day sampling period, this left us with 8,794 vignettes. Using a selection method based on R and Shiny Dashboard, we randomly drew one vignette at a time and manually either included it in the relevant quota or excluded it based on our predefined exclusion criteria. When a specific quota was filled, any new vignettes in that category were excluded. We continued this procedure until all quotas were filled. Once we compiled our vignette set, we made minor adjustments, such as correcting typographical errors and standardizing units of measurement (e.g., adding “kg” to vignettes that originally reported weights in “lbs”). We did not edit the vignettes in any other way, because the principles of representative design require stimuli to be as close to the actual decision environment as possible. Any modification could unpredictably affect the cues and their correlations to triage decisions and thus the representativeness of the vignettes. Using this sampling method, the RepVig Framework, we compiled a final set of 45 vignettes to construct a set that is comparable in size to previous studies^10,14^.

In alignment with established best practices and previous SAA research^14,16,39^, two licensed physicians assessed the appropriate triage level of each vignette independently. After rating all vignettes, the physicians discussed their answers in cases of disagreement to reach consensus on the appropriate triage level, thereby finalizing the solutions for this set. This approach guarantees that the triage levels assigned to each vignette are not only based on expert medical opinion but also on a harmonized understanding between the two assessing physicians^14,16,39^.

For the evaluation study, we compared data collected with our novel sampling approach to data collected with vignettes developed by Semigran et al.^10^. This dataset was selected for comparison, because it is openly accessible and has been used to evaluate the capabilities of both medical laypeople and SAAs. The dataset focusing on laypeople’s capability^34^ was published in 2021 and comprises data from 91 participants, each of whom rated all 45 vignettes developed by Semigran et al. The dataset assessing SAA accuracy^35^ replicated an earlier accuracy study by utilizing the same set of vignettes and SAA selection criteria. The authors tested 22 different SAAs using the same set of 45 vignettes from Semigran et al. that were used in the study examining laypeople’s capabilities.

### Data Collection

To compare triage performance with both vignette sets, we collected data from three different groups: Medical laypeople, SAAs and LLMs. To obtain data from laypeople, we used Prolific, an online panel provider known for its high-quality data^40^. We drew a random sample from this platform, including participants from the United States (to ensure comparability with our comparator dataset) and excluding participants that were health professionals. Based on our comparator dataset^41^, we aimed to get about 4,000 ratings to arrive at a similar number of ratings as in the comparator dataset (4,095 ratings) to ensure comparability and adequate statistical power to detect effects of similar size under similar conditions. To ensure attentive participation, each person was assigned a random sample of 20 out of the 45 vignettes.

Thus, we collected data from 202 participants between the October 18^th^ and 19^th^ 2023 using an online survey designed with Unipark (Questback)^42^. Based on Levine et al.’s easy-to-comprehend and laypeople-friendly phrasing of different urgency levels^12^, participants were asked to choose whether the most appropriate level for each vignette is emergency care, urgent care, non-emergent care or self-care. To further motivate patients, we offered a bonus of 0.02$ for each correctly solved vignette, in addition to a base remuneration of 1$ for 10 minutes. Additionally, we embedded an attention check in two fictitious vignettes, instructing participants to select a specific response. Data from participants who failed to pass at least one attention check were excluded from the analysis.

Following best practices to reduce effects from inputter variability, data from SAAs were collected by two graduate students (DS and SZ) – without professional medical training – who entered the cases into different SAAs between October 24^th^ 2023 and January 22^nd^ 2024. Having students enter the vignettes rather than health professionals ensured that only information stated in the vignettes were entered without subjective clinical interpretation of symptoms, which could introduce bias^17^. The two students entered each vignette into 12 different SAAs (amounting to 540 assessments). The selection of SAAs was based on our comparator dataset^13^ to ensure comparability. The original study from which this dataset is derived had systematically identified all publicly available SAAs. For our study, we had to exclude certain SAAs because four were no longer available, two discontinued providing triage advice, two had shifted to a subscription model, and two now give specialized advice for pediatric cases. A comprehensive list of the remaining 12 SAAs is provided in the Appendix. Inputters were blinded to the outcome (i.e. the correct solution) and recoded the triage levels as provided by the SAAs. These results were then reviewed by the lead author (MK). In instances where the inputters came up with different results, we considered a vignette correctly solved if at least one of them had recorded the correct solution. This criterion was based on the rationale that if one inputter could derive the correct solution, the SAA was technically capable of providing an accurate triage assessment^43^. Analyses for each inputter separately can be found in the Appendix.

Data from LLMs were collected by one person only between November 28^th^ and 29^th^ 2023, as we used a predefined, one-shot prompt for all LLMs to circumvent input variation. We collected data for both the vignettes from our comparator dataset and those developed in this study. The prompt we used was adapted from two previous studies^12,44^ and stated

> *“Please give a triage classification for the following vignette. The criteria for triage classifications are presented below: Emergency: Call 911 or go directly to the emergency room; Non-Emergency: Try to see a doctor, but the symptoms do not require immediate attention at an emergency room. For example, you can see a doctor in the next week. Self-care: Let the health issue get better on its own and review the situation in a few days again*.*”*.

We tested all LLMs that we could identify through a systematic search (entering combinations of the words “large language model”, “LLM”, “chatbot”, “GPT”, “text generation” and “BERT” into search engines) and that offered a web interface. In total, we evaluated five LLMs. The list of LLMs can be found in the Appendix.

### Data Analysis

To align our study with the comparator datasets, which did not include ‘urgent care’ as a distinct solution, we recoded all ‘urgent care’ responses to ‘non-emergency care’. This adjustment ensures consistency in our analysis, as ‘urgent care’ was originally classified as ‘non-emergency care’ in the comparator datasets.

We then proceeded to compare the entire set of vignettes obtained through our novel sampling approach with the vignette set commonly used in previous studies assessing SAA accuracy^10^. For the analysis involving medical laypeople, SAAs and LLMs, we employed both descriptive and inferential statistical methods. To examine the differences between our newly sampled vignette set and the traditional set used for laypeople and SAAs, we used a mixed-effects binomial logistic regression. In these models, participants (or SAAs or LLMs) were treated as a random effect, while the vignette sets were considered a fixed effect.

Our study’s outcome measures were aligned with established reporting guidelines^36^ and included overall accuracy, accuracy for each self-triage level separately, the safety of advice (calculated as the percentage of emergencies identified), the comprehensiveness (how many of the total number of vignettes received a solution), and the inclination to overtriage (the percentage of overtriage errors among all errors). Since not all SAAs gave advice for each vignette, we calculated a Capability Comparison Score (CCS, as elaborated in Kopka et al.^36^). This metric adjusts the accuracy scores to reflect the varying difficulty levels of vignettes that different SAAs were able to address, allowing for a more equitable comparison of capability across SAAs that may not have provided solutions for all cases. For assessing the interrater agreement on SAA data, we used two-way mixed, agreement, average-measures intra-class correlation (ICC), as solutions were coded as ordinal^45^. Values above .40 were considered acceptable^46^.

## Results

### Vignette Sampling

The vignette sampling process is displayed in Figure 2 based on the PRISMA reporting standard^47^. Over a 14-day period, we identified 8,794 posts, of which 858 were duplicates. After removing cases that were too long, we arrived at a final set of 5,388 posts. We randomly sampled from these posts until all quotas were filled, reviewing 526 vignettes in total. Out of these, 423 were excluded based on the exclusion criteria and 58 because quota limits were reached.

**Figure 2.**
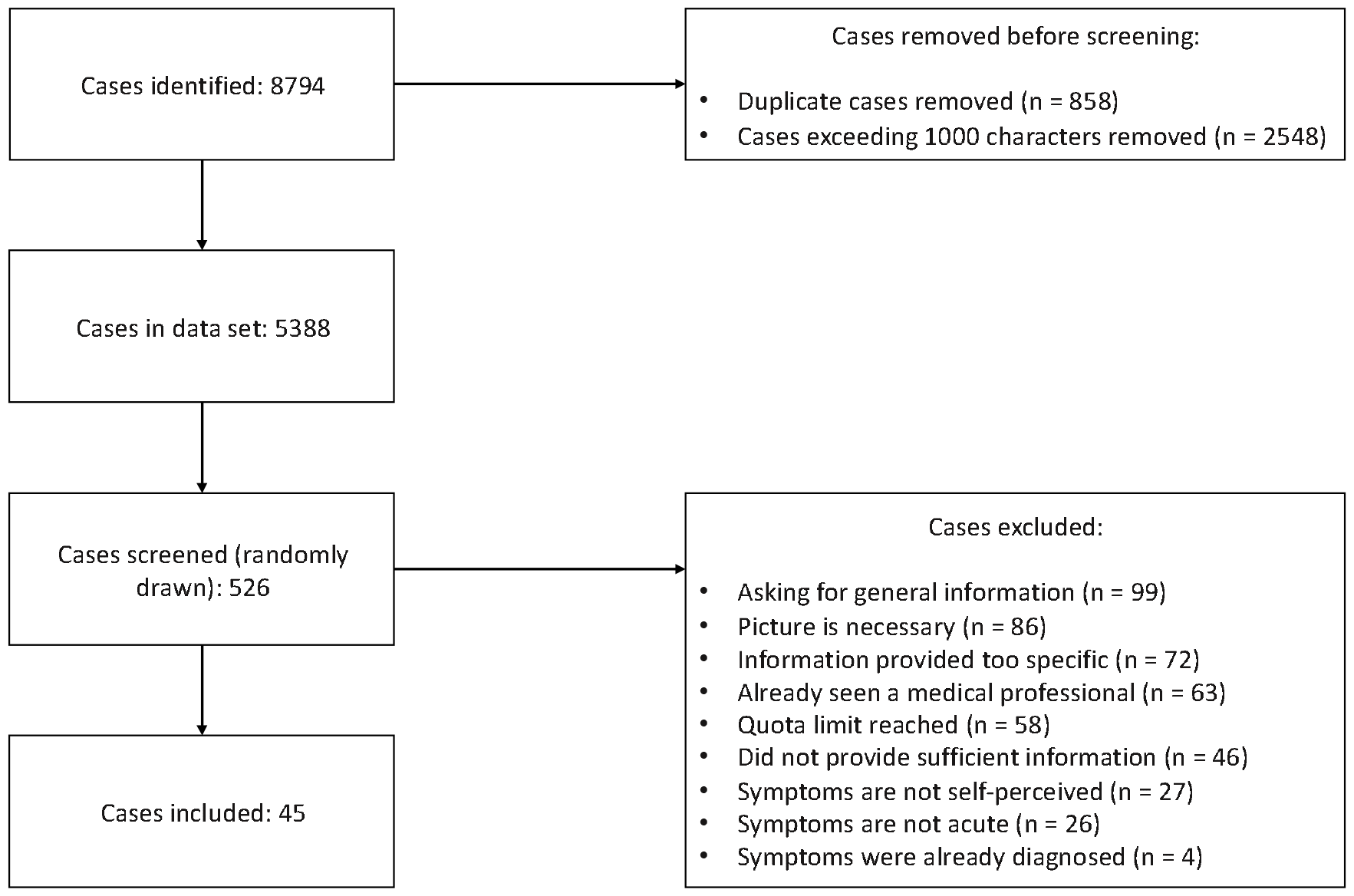
PRISMA chart with number of case vignettes identified, screened, excluded and included.

### Participants

Out of the 33,736 eligible participants on the Prolific platform, 204 participants started the survey and 202 completed it. Data from four participants were excluded, because they failed at least one attention check. This resulted in a final sample size of 198 participants with 20 assessments each and a total number of 3,960 vignette assessments. Descriptions of participants’ characteristics can be found in Table 2.

**Table 2.** Description of participants. N = 198, M = Mean, SD = Standard Deviation, n = number.

| Characteristic | Result |
| --- | --- |
| **Age, M (SD)** | 40.70 (13.85) |
| **Gender, n (%)** |  |
| Male | 97 (48.99%) |
| Female | 98 (49.49%) |
| Other | 3 (1.52%) |
| **Education, n (%)** |  |
| Less than high school diploma | 2 (1.01%) |
| High school graduate, GED, or alternative | 29 (14.46%) |
| Some college or Associate degree | 61 (30.81%) |
| Bachelor degree | 73 (36.87%) |
| Graduate degree or higher | 33 (16.67%) |
| **Medical training, n (%)** |  |
| No training at all | 165 (83.33%) |
| Basic first aid training | 33 (16.67%) |
| **eHealth Literacy** (quantified by the eHealth Literacy Scale), M (SD) | 30.4 (4.37) |

Self-Triage Performance Metrics Laypeople’s Performance

On average, there was no performance difference for laypeople triaging vignettes sampled for this study and vignettes that were traditionally used (OR = 0.94, SE = 0.05, z = -1.40, p. = 0.16). However, laypeople identified emergency cases more often using representative vignettes compared to traditional vignettes (OR = 1.77, SE = 0.21, z = 2.72, p = 0.006). The same pattern emerged for non-emergency cases (OR = 1.27, SE = 0.08, z = 2.94, p = 0.003). Conversely, laypeople identified self-care cases less often with representative than with traditional vignettes (OR = 0.59, SE = 0.10, z = -5.08, p < .001). As a result, their judgement based on representative vignettes can be considered safer than based on traditional vignettes (OR = 1.84, SE = 0.08, z = 7.38, p < 0.001). Overall, laypeople were more inclined to overtriage with representative vignettes compare to traditional vignettes (OR = 2.10, SE = 0.10, z = 7.43, p < .001). A summary of these findings can be found in Table 3.

**Table 3.**
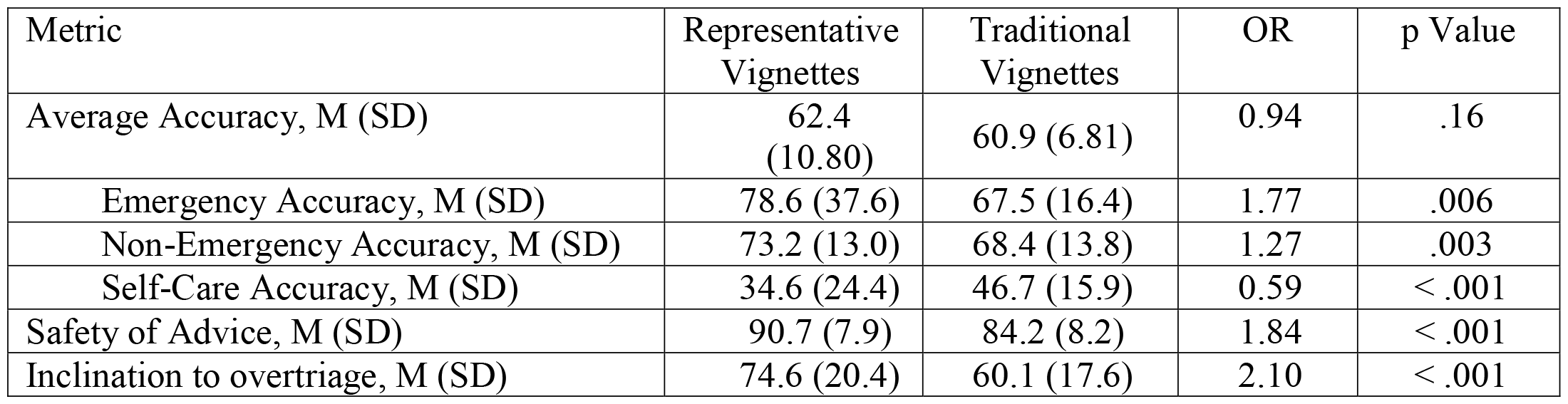
Laypeople’s self-triage performance with representative versus traditional vignettes, M = mean, SD = standard deviation.

| Metric | Representative Vignettes | Traditional Vignettes | OR | p Value |
| --- | --- | --- | --- | --- |
| Average Accuracy, M (SD) | 62.4 (10.80) | 60.9 (6.81) | 0.94 | .16 |
| Emergency Accuracy, M (SD) | 78.6 (37.6) | 67.5 (16.4) | 1.77 | .006 |
| Non-Emergency Accuracy, M (SD) | 73.2 (13.0) | 68.4 (13.8) | 1.27 | .003 |
| Self-Care Accuracy, M (SD) | 34.6 (24.4) | 46.7 (15.9) | 0.59 | < .001 |
| Safety of Advice, M (SD) | 90.7 (7.9) | 84.2 (8.2) | 1.84 | < .001 |
| Inclination to overtriage, M (SD) | 74.6 (20.4) | 60.1 (17.6) | 2.10 | < .001 |

### SAA Performance

Agreement between both SAA inputters was acceptable (ICC = 0.605). Across all SAAs, triage accuracy was significantly higher for representative vignettes compared to traditional vignettes (OR = 2.00, SE = 0.139, z = 5.68, p < .001). In detecting emergencies, we could not find a statistically significant difference (OR = 2.26, SE = 0.53, z = 1.52, p = 0.13), but SAAs detected non-emergency cases (OR = 2.38, SE = 0.236, z = 3.70, p < .001) and self-care cases more often (OR = 2.53, SE = 0.292, z = 3.18, p = .0015) in representative vignettes compared to traditional vignettes. The safety of advice was higher with representative vignettes for one inputter (OR = 1.81, SE = 0.224, z = 2.64, p = .008), and we found a similar but non-significant trend for the second inputter (OR = 1.42, SE = 0.206, z = 1.70, p = .09). With representative vignettes, the inclination to overtriage was higher for one inputter (OR = 1.80, SE = 0.277, z = 2.112, p = .035) and lower for the other inputter (OR = 0.53, SE = 0.277, z = -2.30, p = .022). See Table 4 for a summary.

**Table 4.** SAA’s self-triage performance with representative versus traditional vignettes, M = mean, SD = standard deviation.

| Metric | Representative Vignettes | Traditional Vignettes | OR | p Value |
| --- | --- | --- | --- | --- |
| Average Accuracy, M (SD) | 67.8 (46.8) | 48.9 (50.0) | 2.00 | < .001 |
| Emergency Accuracy, M (SD) | 75.0 (44.2) | 54.4 (49.9) | 2.26 | .013 |
| Non-Emergency Accuracy, M (SD) | 76.4 (42.5) | 60.6 (49.0) | 2.38 | < .001 |
| Self-Care Accuracy, M (SD) | 46.8 (50.1) | 31.7 (46.6) | 2.53 | .002 |
| Safety of Advice*, M (SD) | 83.7 (11.1)<br>92.2 (5.3) | 85.6 (8.1) | 1.81 / 1.42 | .008 / .09 |
| Inclination to overtriage*, M (SD) | 52.5 (29.0)<br>74.0 (17.6) | 56.6 (23.8) | 1.80 / 0.53 | .035 / .022 |

\* Since no aggregated statistic can be calculated for this metric, values for both inputters are reported.

The average item difficulty for representative vignettes among SAAs was higher (M = 0.68, SD = 0.21) than for the traditional vignettes (M = 0.49, SD = 0.26). Performance comparisons with CCS values and ranks are summarized in Table 5. Detailed metrics for every individual SAA can be found in the Appendix.

**Table 5.**
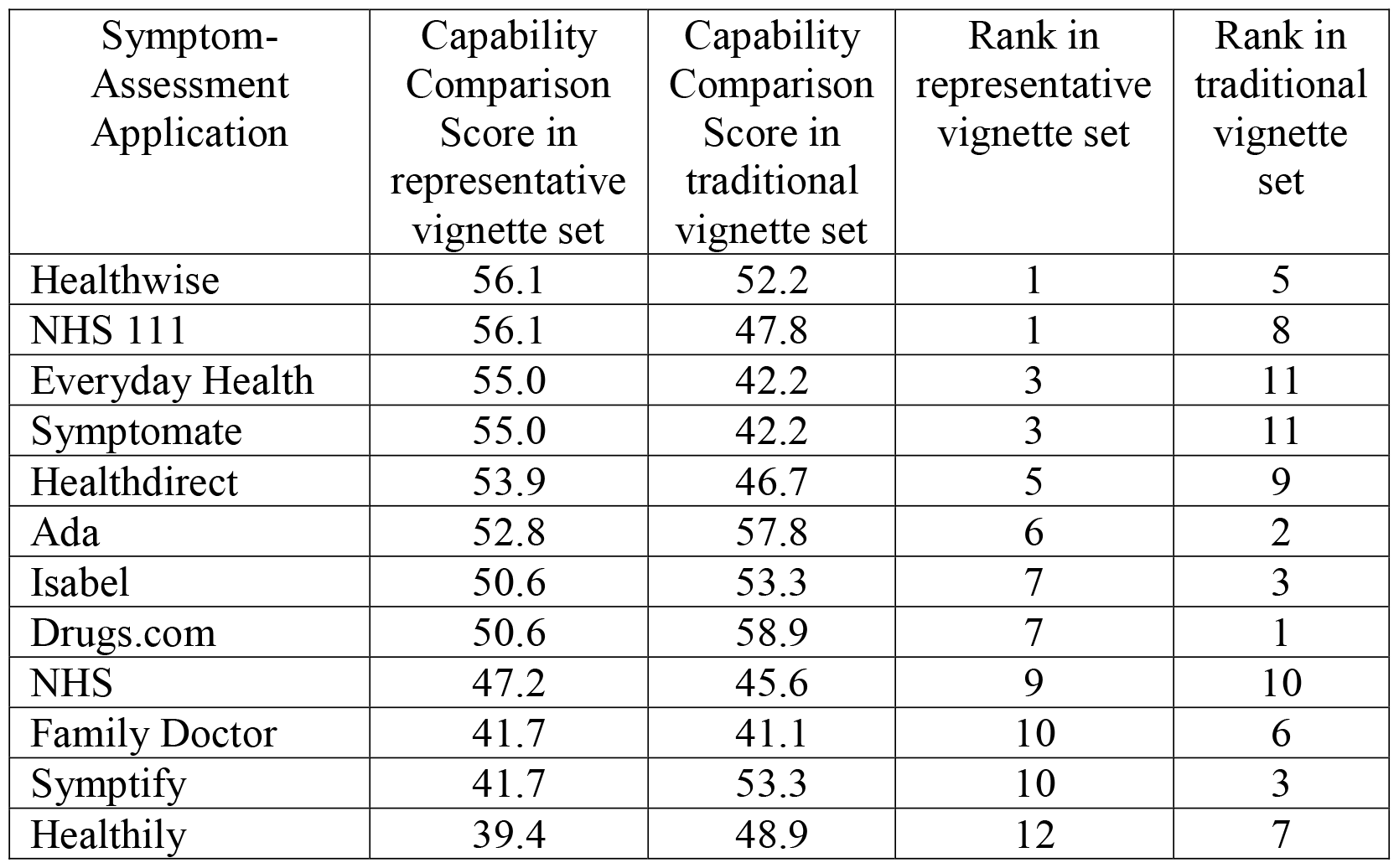
Capability comparison scores (CCS) and ranks for SAAs in both vignette sets

### LLM Performance

Across all LLMs, triage accuracy was significantly higher for representative vignettes compared to traditional vignettes (OR = 1.52, SE = 0.196, z = 2.14, p = .03). In detecting emergencies, we could not find a statistically significant difference (OR = 0.37, SE = 0.80, z = -1.25, p = .21), but LLMs detected non-emergency cases more often (OR = 3.01, SE = 0.00, z = 358.9, p < .001) and self-care cases less often (OR = 0.25, SE = 0.00, z = -449.8, p < .001) in representative compared to traditional vignettes. The safety of advice from LLMs was higher with representative vignettes compared to traditional vignettes (OR = 3.41, SE = 0.39, z = 3.19, p = .001). Overall, LLMs were more likely to overtriage with representative vignettes (OR = 2.66, SE = 0.43, z = 2.30, p = 0.022). A summary can be found in Table 6.

**Table 6.** LLM’s self-triage performance with representative versus traditional vignettes, M = mean, SD = standard deviation.

| Metric | Representative Vignettes | Traditional Vignettes | OR | p Value |
| --- | --- | --- | --- | --- |
| Average Accuracy, M (SD) | 67.6 (2.5) | 57.8 (9.4) | 1.52 | .03 |
| Emergency Accuracy, M (SD) | 50.0 (35.4) | 66.7 (36.5) | 0.37 | .21 |
| Non-Emergency Accuracy, M (SD) | 95.3 (3.8) | 88.0 (17.9) | 3.01 | < .001 |
| Self-Care Accuracy, M (SD) | 6.15 (10.0) | 18.7 (17.3) | 0.25 | < .001 |
| Safety of Advice, M (SD) | 95.6 (3.5) | 87.1 (14.5) | 3.41 | .001 |
| Inclination to overtriage, M (SD) | 86.5 (11.0) | 73.6 (23.5) | 2.66 | .022 |

Because all LLMs provided solutions for all vignettes, all LLMs were tested with vignettes of the same item difficulty. For representative vignettes the average item difficulty was M = 0.68 (SD = 0.42) and for traditional vignettes the average item difficulty was M = 0.58 (SD = 0.34). CCS values and ranks for each LLM are summarized in Table 7.

**Table 7.** Capability comparison scores (CCS) and ranks for LLMs in both vignette sets

| Large Language Model | Capability Comparison Score in representative vignette set | Capability Comparison Score in traditional vignette set | Rank in representative vignette set | Rank in traditional vignette set |
| --- | --- | --- | --- | --- |
| GPT-4 (ChatGPT) | 51.8% | 53.3% | 1 | 1 |
| Llama 2 | 50.7% | 42.2% | 2 | 5 |
| PaLM 2 (Google Bard) | 49.6% | 48.9% | 3 | 4 |
| Pi | 49.6% | 53.3% | 4 | 2 |
| Claude 2 | 48.4% | 52.2% | 5 | 3 |

## Discussion

In this study, we aimed to extend Brunswik’s representative design principles to self-triage audit studies and used this approach to generate a new set of vignettes for studying self-triage performance of laypeople, SAAs, and LLMs through a representative design approach and the RepVig Framework. An extensive pool of case descriptions was sampled in the target environment and provided a robust basis for randomly selecting cases, which were then stratified by symptom clusters to obtain a final set of vignettes that is representative also in terms of the distribution of cases. Then, we examined whether these vignettes result in different performance patterns than traditional vignettes. That was the case for all agents we tested, that is, for laypeople, SAAs, and LLMs, which we discuss in turn.

Although average accuracy of laypeople was similar for both vignette sets, they were more accurate for emergencies and non-emergencies, and less accurate for self-care cases. The safety of advice was also significantly higher (albeit with more overtriage errors, i.e., rating symptoms as more urgent than they are). From an ecological rationality perspective, these findings make sense: because individuals face no harm in calling a general practitioner but might face negative health outcomes if they experience symptoms that require treatment but do not seek health care, they can be expected to be risk averse and seek care rather than stay at home. Unlike a previous study, which used traditional vignettes and reported that laypeople have difficulty identifying emergencies^48^, our results based on representative vignettes suggest that laypeople identify emergencies more reliably and have difficulty identifying self-care cases. Thus, from an applied perspective, decision support tools should focus on helping laypeople identify self-care cases rather than emergencies.

For SAAs, we observed higher accuracy overall and for each triage level when using representative vignettes. However, we also found a slightly lower safety of advice compared to the traditional set, whereas our findings regarding inclination to overtriage are inconclusive. When comparing different SAAs using the CCS, rankings varied significantly between traditional and representative vignettes. This highlights the crucial role of the vignette sets in SAA testing. When selecting an SAA for general use in guiding patients through the healthcare system, these different vignettes would lead to varying choices.

In LLMs, we observed significant differences across all estimates when comparing representative and traditional vignette sets. The average accuracy was higher with representative vignettes, but the distribution across different triage levels varied. Emergency and self-care cases were less frequently identified correctly, while non-emergency cases were identified more often. The advice was also safer with representative vignettes, although they led to more overtriage errors. The ranking of LLMs also changed when using representative vignettes.

Overall, we found that our results differed when using representative vignettes, aligning with our hypothesis and findings from several other empirical studies^25,27,29,30^. This suggests that employing a representative design approach is valuable for evaluating laypeople’s self-triage performance with and without SAAs. Importantly, we do not claim that the sampling method presented in this article is or should be the only and best way to evaluate SAAs. On the contrary, in line with the concept of representative design, we suggest that each use case requires its specific set of stimuli (e.g., case vignettes sampled from different populations in specific situations). In this sense, also traditional vignettes can provide valuable insights, for instance, to examine how SAAs might perform when being used by *clinicians*. Similarly, studying ED cases (as done by Fraser et al.^49^) could help determine if patients visiting the ED might have opted for primary care had they consulted an SAA beforehand. With a sample of primary care cases, the potential effect of SAA on the redistribution of care seeking efforts could be assessed from a systems perspective.

Therefore, we suggest that future self-triage evaluation studies should specify to which particular decision situation they wish to generalize and sample case vignettes from that situation instead of relying on one fixed set of vignettes independent of the use case at hand. This recommendation is especially relevant considering that LLMs (and perhaps SAAs in the future) – that are regularly updated with new data^50^ and expected to significantly impact future healthcare^51,52^ – might be trained on published vignettes or could retrieve results for these vignettes through web searches. Thus, generalizability for LLMs is limited because published vignettes and their solutions can be part of the training set, and a standardized approach to vignette sampling (without a published solution) and continuously generating new vignettes may be more beneficial than using a fixed set of vignettes. Interestingly, by using more specific vignettes, researchers will ultimately provide more generalizable insights for clearly defined use cases.

The approach presented in this article is particularly pertinent to achieving the general goal of SAAs, which is to enable people to assess symptoms without a clinician’s direct involvement. Beyond that use case, we propose our RepVig Framework based on the representative design approach as a standard for sampling vignettes from situations to which one wishes to generalize. For regulators evaluating SAAs and other digital health technologies, our study emphasizes the importance of data quality. Testing decision support systems should be done both with relevant metrics^36^ *and* with reliable vignettes that allow generalizations beyond the testing scenario. Because using unrepresentative vignettes for system testing can significantly influence the results – potentially leading to biased outcomes – only using representative vignettes and providing information on the vignettes that SAAs are tested with is particularly important. Stakeholders should specify the population they want to generalize to and use corresponding vignettes for evaluations.

This study has several limitations. First, we could not obtain additional information for these cases. Although this is also a common issue with traditional vignettes, having extra details available that SAAs might inquire about would be beneficial. This way, inputters get more precise results because they are able to respond to SAA questions throughout the interaction. However, interviewing individuals in the situation of experiencing the symptoms to get additional information poses significant challenges; it is particularly unfeasible and unethical for those with urgent symptoms. Therefore, this trade-off between realism and practical and ethical considerations we propose is likely the most acceptable balance achievable. A second limitation pertains to the results obtained from LLMs. These results are based on a specific prompt, and real-world interaction might yield different results due to varying prompts, interactions, and interpretation of LLM outputs. Additionally, LLM output is non-deterministic^53^, meaning results could vary even if cases are entered using the same prompt again. Given rapid improvement cycles, the accuracy of LLMs might change quickly and require ongoing tests. Lastly, the observed disparities in the SAAs’ inclination to overtriage are likely due to inputter variability, as it has been previously shown to affect such estimates^43^. However, we followed standard practice in symptom checker audit studies and this variance is present in similar studies as well. While technically all differences might stem from inputter variability, our results for laypeople and LLMs – in which inputter variability is impossible – differ between these two vignette sets too. Therefore, we believe that our results are not solely attributable to inputter variability.

In summary, we propose refining vignettes in self-triage studies to enhance generalizability of the observed performance patterns. To ground the vignette sampling process on a theoretical basis, we suggest using representative design principles for investigating self-triage of laypeople (see Figure 1) and the RepVig Framework as a general framework for sampling case vignettes and evaluating triage performance in all kinds of use cases.

## Data Availability

The data will be published in an open access repositorium upon acceptance.

## Data availability

The data will be published in an open access repositorium upon acceptance.

## Author contributions

MK and MAF conceived of the study. MK developed the methodology with critical input from MAF, created the vignettes and handled programming and sampling. HN and MP provided solutions to the vignettes. MK collected the data, with DS and SZ supporting data collection from symptom-assessment applications. MK conducted the analyses and data visualization and wrote the first draft of the manuscript. All authors provided critical input and worked on manuscript development.

## References

1. Napierala, H. et al. Examining the impact of a symptom assessment application on patient-physician interaction among self-referred walk-in patients in the emergency department (AKUSYM): study protocol for a multi-center, randomized controlled, parallel-group superiority trial. Trials 23, 791 (2022).

2. Kopka, M. et al. Characteristics of Users and Nonusers of Symptom Checkers in Germany: Cross-Sectional Survey Study. J Med Internet Res 25, e46231 (2023).

3. Aboueid, S., Meyer, S. B., Wallace, J. & Chaurasia, A. Latent classes associated with the intention to use a symptom checker for self-triage. PLoS ONE 16, e0259547 (2021).

4. Aboueid, S., Meyer, S., Wallace, J. R., Mahajan, S. & Chaurasia, A. Young Adults’ Perspectives on the Use of Symptom Checkers for Self-Triage and Self-Diagnosis: Qualitative Study. JMIR Public Health Surveill 7, e22637 (2021).

5. Kopka, M. et al. Determinants of Laypersons’ Trust in Medical Decision Aids: Randomized Controlled Trial. JMIR Hum Factors 9, e35219 (2022).

6. Gellert, G. A. et al. A multinational survey of patient utilization of and value conveyed through virtual symptom triage and healthcare referral. Front. Public Health 10, 1047291 (2023).

7. Morse, K. E., Ostberg, N. P., Jones, V. G. & Chan, A. S. Use Characteristics and Triage Acuity of a Digital Symptom Checker in a Large Integrated Health System: Population-Based Descriptive Study. Journal of Medical Internet Research 22, e20549 (2020).

8. Wallace, W. et al. The diagnostic and triage accuracy of digital and online symptom checker tools: a systematic review. npj Digit. Med. 5, 118 (2022).

9. Riboli-Sasco, E. et al. Triage and Diagnostic Accuracy of Online Symptom Checkers: Systematic Review. J Med Internet Res 25, e43803 (2023).

10. Semigran, H. L., Linder, J. A., Gidengil, C. & Mehrotra, A. Evaluation of Symptom Checkers for Self Diagnosis and Triage: Audit Study. BMJ 351, 1–9 (2015).

11. Semigran, H. L., Levine, D. M., Nundy, S. & Mehrotra, A. Comparison of Physician and Computer Diagnostic Accuracy. JAMA Internal Medicine 176, 1860–1861 (2016).

12. Levine, D. M. et al. The Diagnostic and Triage Accuracy of the GPT-3 Artificial Intelligence Model. http://medrxiv.org/lookup/doi/10.1101/2023.01.30.23285067 (2023) xdoi:10.1101/2023.01.30.23285067.

13. Schmieding, M. L. et al. Triage Accuracy of Symptom Checker Apps: 5-Year Follow-up Evaluation. J Med Internet Res 24, e31810 (2022).

14. Hill, M. G., Sim, M. & Mills, B. The Quality of Diagnosis and Triage Advice Provided by Free Online Symptom Checkers and Apps in Australia. Med J Aust 212, 514–519 (2020).

15. Ceney, A. et al. Accuracy of online symptom checkers and the potential impact on service utilisation. medRxiv 2020.07.07.20147975 (2020) doi:10.1101/2020.07.07.20147975.

16. Delshad, S., Dontaraju, V. S. & Chengat, V. Artificial Intelligence-Based Application Provides Accurate Medical Triage Advice When Compared to Consensus Decisions of Healthcare Providers. Cureus (2021) doi:10.7759/cureus.16956.

17. El-Osta, A. et al. What is the suitability of clinical vignettes in benchmarking the performance of online symptom checkers? An audit study. BMJ Open 12, e053566 (2022).

18. Painter, A., Hayhoe, B., Riboli-Sasco, E. & El-Osta, A. Online Symptom Checkers: Recommendations for a Vignette-Based Clinical Evaluation Standard. J Med Internet Res 24, e37408 (2022).

19. Yu, S. W. Y. et al. Triage Accuracy of Online Symptom Checkers for Accident and Emergency Department Patients. Hong Kong Journal of Emergency Medicine 27, 217–222 (2020).

20. Berry, A. C. et al. Online symptom checker diagnostic and triage accuracy for HIV and hepatitis C. Epidemiol. Infect. 147, e104 (2019).

21. Dhami, M. K., Hertwig, R. & Hoffrage, U. The Role of Representative Design in an Ecological Approach to Cognition. Psychological Bulletin 130, 959–988 (2004).

22. Meyer, A. N. D. et al. Patient Perspectives on the Usefulness of an Artificial Intelligence-Assisted Symptom Checker: Cross-Sectional Survey Study. J. Med. Internet Res. 22, e14679 (2020).

23. Brunswik, E. Representative design and probabilistic theory in a functional psychology. Psychological Review 62, 193–217 (1955).

24. Waghorn, J. et al. Clinical Judgement Analysis: An innovative approach to explore the individual decision-making processes of pharmacists. Research in Social and Administrative Pharmacy 17, 2097– 2107 (2021).

25. Cesario, J. What can experimental studies of bias tell us about real-world group disparities? Behav Brain Sci 45, e66 (2022).

26. Richter, B. & Hütter, M. Learning of affective meaning: revealing effects of stimulus pairing and stimulus exposure. Cognition and Emotion 35, 1588–1606 (2021).

27. Fritzsche, B. A. & Brannick, M. T. The importance of representative design in judgment tasks: The case of résumé screening. J Occupat & Organ Psyc 75, 163–169 (2002).

28. Miller, L. C., Wang, L., Jeong, D. C. & Gillig, T. K. Bringing the Real World into the Experimental Lab: Technology-Enabling Transformative Designs. in Social-Behavioral Modeling for Complex Systems 359–386 (John Wiley & Sons, Ltd, 2019). doi:10.1002/9781119485001.ch16.

29. Wang, X. & Navarro-Martinez, D. Bridging the gap between the economics lab and the field: Dictator games and donations. Judgm. decis. mak. 18, e18 (2023).

30. Gigerenzer, G., Hoffrage, U. & Kleinbölting, H. Probabilistic mental models: A Brunswikian theory of confidence. Psychological Review 98, 506–528 (1991).

31. Karelaia, N. & Hogarth, R. M. Determinants of linear judgment: A meta-analysis of lens model studies. Psychological Bulletin 134, 404–426 (2008).

32. Ayers, J. W. et al. Comparing Physician and Artificial Intelligence Chatbot Responses to Patient Questions Posted to a Public Social Media Forum. JAMA Intern Med (2023) doi:10.1001/jamainternmed.2023.1838.

33. Von Elm, E. et al. The Strengthening the Reporting of Observational Studies in Epidemiology (STROBE) statement: guidelines for reporting observational studies. The Lancet 370, 1453–1457 (2007).

34. Schmieding, M. L., Mörgeli, R., Schmieding, M. A. L., Feufel, M. A. & Balzer, F. Data set supplementing ‘Benchmarking triage capability of symptom checkers against that of medical laypersons: Survey study’. Zenodo 10.5281/ZENODO.4454537 (2021).

35. Schmieding, M. L. et al. Data Set on Accuracy of Symptom Checker Apps in 2020. Zenodo 10.5281/ZENODO.6054092 (2022).

36. Kopka, M., Feufel, M. A., Berner, E. S. & Schmieding, M. L. How suitable are clinical vignettes for the evaluation of symptom checker apps? A test theoretical perspective. DIGITAL HEALTH 9, 20552076231194929 (2023).

37. Kopka, M. & Feufel, M. A. symptomcheckR: An R Package for Analyzing and Visualizing Symptom Checker Performance. http://medrxiv.org/lookup/doi/10.1101/2024.02.06.24302384 (2024) xdoi:10.1101/2024.02.06.24302384.

38. Arellano Carmona, K., Chittamuru, D., Kravitz, R. L., Ramondt, S. & Ramírez, A. S. Health Information Seeking From an Intelligent Web-Based Symptom Checker: Cross-sectional Questionnaire Study. J Med Internet Res 24, e36322 (2022).

39. Gilbert, S. et al. How accurate are digital symptom assessment apps for suggesting conditions and urgency advice? A clinical vignettes comparison to GPs. BMJ Open 10, e040269 (2020).

40. Peer, E., Rothschild, D. M., Evernden, Z., Gordon, A. & Damer, E. MTurk, Prolific or Panels? Choosing the Right Audience for Online Research. SSRN Journal (2021) doi:10.2139/ssrn.3765448.

41. Schmieding, M. L., Mörgeli, R., Schmieding, M. A. L., Feufel, M. A. & Balzer, F. Benchmarking Triage Capability of Symptom Checkers Against That of Medical Laypersons: Survey Study. J Med Internet Res 23, e24475 (2021).

42. Questback GmbH. Umfragesoftware für Studierende und Wissenschaftler. Unipark https://www.unipark.com/umfragesoftware/ (2021).

43. Meczner, A. et al. Accuracy as a Composite Measure for the Assessment of Online Symptom Checkers in Vignette Studies: Evaluation of Current Practice and Recommendations (Preprint). http://preprints.jmir.org/preprint/49907 (2023) xdoi:10.2196/preprints.49907.

44. Ito, N. et al. The Accuracy and Potential Racial and Ethnic Biases of GPT-4 in the Diagnosis and Triage of Health Conditions: Evaluation Study. JMIR Med Educ 9, e47532 (2023).

45. Hallgren, K. A. Computing Inter-Rater Reliability for Observational Data: An Overview and Tutorial. TQMP 8, 23–34 (2012).

46. Cicchetti, D. V. Guidelines, criteria, and rules of thumb for evaluating normed and standardized assessment instruments in psychology. Psychological Assessment 6, 284–290 (1994).

47. Page, M. J. et al. The PRISMA 2020 statement: an updated guideline for reporting systematic reviews. Syst Rev 10, 89 (2021).

48. Kopka, M., Feufel, M. A., Balzer, F. & Schmieding, M. L. The Triage Capability of Laypersons: Retrospective Exploratory Analysis. JMIR Form Res 6, e38977 (2022).

49. Fraser, H. S. F. et al. Evaluation of Diagnostic and Triage Accuracy and Usability of a Symptom Checker in an Emergency Department: Observational Study. JMIR Mhealth Uhealth 10, e38364 (2022).

50. Jang, J. et al. Towards Continual Knowledge Learning of Language Models. (2021) doi:10.48550/ARXIV.2110.03215.

51. Meskó, B. & Topol, E. J. The imperative for regulatory oversight of large language models (or generative AI) in healthcare. npj Digit. Med. 6, 120 (2023).

52. Thirunavukarasu, A. J. et al. Large language models in medicine. Nat Med 29, 1930–1940 (2023).

53. Lee, M., Liang, P. & Yang, Q. CoAuthor: Designing a Human-AI Collaborative Writing Dataset for Exploring Language Model Capabilities. in CHI Conference on Human Factors in Computing Systems 1–19 (ACM, New Orleans LA USA, 2022). doi:10.1145/3491102.3502030.

